# Enhancing Blood Availability in Latin America: A Study on Public Perceptions and Barriers to Blood Donation in Guatemala during the COVID-19 Pandemic

**DOI:** 10.1101/2024.06.16.24309008

**Authors:** Carolina Torres Perez-Iglesias, Jose C. Monzon, Isabella Faria, Shreenik Kundu, Ahsan Zil-E-Ali, Rashi Jhunjhunwala, Nakul Raykar, Sabrina Asturias

**Author notes:** Contributions: conceptualization, data curation, formal analysis, investigation, methodology, project administration, resources, supervision, validation, visualization, writing – original draft preparation, writing - review and editing. Contributions: conceptualization, data curation, investigation, project administration, resources, validation, writing – original draft preparation, writing - review and editing. Contributions: data curation, formal analysis, investigation, methodology, writing – original draft preparation, writing - review and editing. Contributions: data curation, formal analysis, investigation, methodology, project administration, writing – review and editing. Contributions: data curation, formal analysis, investigation, methodology, software, writing – original draft preparation. Contributions: writing - review and editing. Contributions: conceptualization, data curation, investigation, methodology, project administration, supervision, validation, visualization, writing - review and editing. Contributions: conceptualization, data curation, investigation, methodology, project administration, resources, supervision, validation, writing - review and editing.

## Abstract

**Objectives:** Guatemala faces a significant challenge with one of the lowest voluntary blood donation rates in Latin America, a problem further exacerbated by the COVID-19 pandemic. This study aimed to investigate the population factors influencing blood donation behavior in Guatemala during the COVID-19 pandemic.

**Methods:** Conducted between August and September 2020, this cross-sectional electronic anonymous survey employed purposive sampling. Participants were questioned about their donation history, knowledge of donation processes, preferences, and barriers and motivators for blood donation. Quantitative and qualitative data were collected and analyzed.

Comparative analyses were conducted based on gender, age, and education level. Regression analysis was used to identify predictors of blood donation behavior. Open-ended responses were studied via thematic content analysis.

**Results:** Among the 1141 respondents, 53.5% reported a history of blood donation. Most donations occurred via referred donations to family or friends (78.5%). Factors such as male gender, older age, and higher education were associated with previous blood donation.

Familiarity with donation centers and willingness to donate strongly influenced donation behavior. Among never donors, 89% expressed willingness to donate. Constraints in accessing donation centers, limited operation hours, insufficient knowledge about the donation process, and concerns over hygiene and safety were identified as the most prevalent barriers to donation.

**Conclusions:** Respondents demonstrated a strong willingness to donate blood voluntarily. Understanding demographic and population factors is critical to inform tailored initiatives to facilitate access to voluntary blood donation in Guatemala.

## INTRODUCTION

Ensuring sufficient and safe access to blood products is a critical component of universal healthcare. Low- and middle-income countries bear a significant proportion of the global burden of diseases, including communicable, maternal-neonatal, nutritional, and traumatic conditions, that often require blood transfusions.^1^ Moreover, these nations commonly grapple with a range of challenges that contribute to the limited availability of blood products, including insufficient integration of blood systems into national health frameworks, disparities in healthcare infrastructure, resource constraints for the collection, processing, and storage of blood products, and persistently low rates of voluntary blood donation. Regrettably, over the last decade, voluntary blood donation rates in Latin America and the Caribbean have experienced a concerning decline,^2^ a trend further exacerbated by the recent COVID-19 pandemic.^3^

Historically, countries in Latin America have depended on replacement-based donation systems, wherein patients must arrange for blood through acquaintances or paid donations rather than relying on voluntary, altruistic contributions.^4,5^ This approach is particularly entrenched due to the region’s diverse demographic and cultural landscape, introducing additional complexities and misconceptions that affect the culture of voluntary blood donation. The COVID-19 pandemic has further amplified concerns regarding the hygiene and safety of blood donation, requiring blood centers to implement enhanced screening protocols to ensure the safety and trust of donors and blood recipients. Given the unmet demand for blood products in the region, there is an urgent need to identify the specific challenges that serve as barriers to voluntary blood donation and to implement targeted strategies to address them.

This study aimed to delve into the factors contributing to the limited availability of blood products in Guatemala, a country with the second-lowest voluntary blood donation rate in Latin America.^6^ Additionally, it emphasizes the profound ramifications of voluntary blood donation for the region’s public health and healthcare equity and highlights the urgent urgency to fortify initiatives to this end.

## METHODS

### Study design

A cross-sectional survey of the population in Guatemala was conducted between August and September 2020 by Dona Guate, a non-profit organization committed to promoting a positive culture of blood donation in Guatemala.^7^ The study aimed to identify public perceptions of voluntary blood donation, discern individual factors that motivate or impede donation and uncover any potential challenges added by the COVID-19 pandemic. The survey was anonymous and administered electronically. It comprised an 11-question instrument in Spanish that featured a combination of multiple-choice, drop-down, and open-ended questions. The translated version of the survey in English can be found in Appendix 1.

### Study distribution

The survey was disseminated through a snowball sampling method through Dona Guate’s social media platform and mailing lists. Participation was entirely voluntary, and no form of compensation was provided. Incomplete responses, individuals under 18, and those residing outside of Guatemala at the time of the survey were excluded from the study.

### Statistical analysis

Statistical analysis was conducted using Stata 16.1 statistical software.^8^ The association between study groups for the categorical variables was assessed using Pearson’s Chi-Square test, which compared the observed and expected responses. The results, absolute percentages, and the count of participants in each category are presented. A multivariate logistic regression model was constructed to identify factors linked with blood donation, including only variables with a p-value <0.05 from the univariate analysis. All the variables analyzed in both the univariate and multivariate regression models were reported with unadjusted and adjusted odd ratios, respectively, along with corresponding 95% confidence intervals. Statistical significance was set at p < 0.05. The qualitative data from the open-ended question underwent thematic content analysis and was reviewed by two independent coders (CT, IF). Frequencies of these identified themes are also presented. This research was approved by the Harvard Longwood Campus Institutional Review Board under protocol IRB 23-0135.

### Ethics statement

This study involved collecting data through an anonymous survey distributed to members of the population. The survey was carefully designed to ensure that no personally identifiable information was collected from participants. As such, the responses could not be traced back to any individual, thereby maintaining complete anonymity.

Given the nature of the data collection and the measures taken to ensure anonymity, the study was deemed to pose minimal risk to participants. The survey focused on gathering non-sensitive information, and there was no potential harm or distress to participants. In accordance with these considerations, it was determined that the study did not meet the criteria for requiring Institutional Review Board approval.

## RESULTS

### Characteristics of Respondents

During the study period, 1141 responses were gathered, primarily from an urban sample of 62.5% (n = 713) of females and 37.5% (n = 428) of males. From these, 94% (n = 1073) resided in Guatemala City at the time of the survey. Two-thirds of the sample were under 40, and over 80% (n = 955) reported a college education or higher. Further demographic details of the respondents are provided in Table 1.

**Table 1.** Comparison of Baseline Characteristics and Responses based on Previous Donation Status.

| Variables | N=1,141 | Has previously donated<br>N = 610 | Has never donated<br>N = 531 | p-value |
| --- | --- | --- | --- | --- |
| **Gender** |  |  |  |  |
| Male | 428 (37.5) | 271 (44.4) | 157 (29.6) | **<0.001** |
| Female | 713 (62.5) | 339 (55.6) | 374 (70.4) |  |
| **Age** |  |  |  |  |
| <=29 | 438 (38.3) | 214 (35.1) | 224 (42.2) | **0.001** |
| 30-39 | 318 (27.9) | 159 (26.1) | 159 (29.9) | 0.172 |
| 40-49 | 151 (13.2) | 86 (14.1) | 65 (12.2) | 0.394 |
| 50-59 | 133 (11.7) | 79 (12.9) | 54 (10.2) | 0.163 |
| 60+ | 101 (8.9) | 72 (11.8) | 29 (5.5) | **<0.001** |
| **Educational level** |  |  |  |  |
| Elementary School/ High School | 186 (16.3) | 91 (14.9) | 95 (17.9) | **0.001** |
| Bachelor | 550 (48.2) | 273 (44.8) | 277 (52.1) |  |
| Master/ Doctorate | 405 (35.5) | 246 (40.3) | 159 (30) |  |
| **Place of residence** |  |  |  |  |
| Guatemala City | 1,073 (94) | 573 (93.9) | 500 (94.2) | 0.907 |
| Other | 68 (6) | 37 (6.1) | 31 (5.8) |  |
| **Do you know where you can donate blood in Guatemala?** |  |  |  |  |
| No | 249 (21.8) | 87 (14.2) | 162 (30.5) | **<0.001** |
| Yes | 892 (78.2) | 523 (85.8) | 369 (69.5) |  |
| **If you were to donate blood, do you care who receives your blood?** |  |  |  |  |
| No | 1,003 (87.9) | 548 (89.8) | 455 (85.7) | **0.032** |
| Yes | 138 (12.1) | 62 (10.2) | 76 (14.3) |  |
| **Are you willing to donate blood?** |  |  |  |  |
| I am not interested in donating blood | 93 (8.2) | 35 (5.7) | 58 (10.9) | **0.001** |
| Yes, but only for emergencies – it does not matter who receives the blood | 256 (22.4) | 135 (22.1) | 121 (22.8) |  |
| Yes, but only for emergencies of family, friends, and people I know | 281 (24.6) | 139 (22.8) | 142 (26.7) |  |
| Yes, I would like to be a frequent donor (1 time per year) | 511 (44.8) | 301 (49.4) | 210 (39.6) |  |

Among the respondents, 53.5% (n = 610) reported a history of blood donation. Additionally, 78.2% (n= 892) reported knowing where to donate blood, and 87.9% (n = 1,003) expressed no preference regarding the recipient of their donated blood. Only 8.2% (n = 93) were not inclined to donate, and 44.8% (n = 511) were interested in becoming frequent donors (at least once annually).

Comparing donor status (previous donors vs. never donors), male respondents exhibited a higher rate of blood donation compared to females (p < 0.001). Respondents under 29 years of age were less commonly previous donors compared to other age groups (p = 0.001), whereas those over 60 were more likely to be previous donors (p < 0.001). Furthermore, a higher education level correlated with a greater likelihood of being a prior blood donor (p = 0.001). Never donors were less likely to know where to donate (p < 0.001) and expressed more concern about who receives the donated blood (p = 0.032).

Among previous donors (n = 610), the most common avenues for donation included referrals to family or friends (77.8%, n = 475), donations via Red Cross campaigns (10%, n = 61), donations via Dona Guate campaigns (3.3% n = 20) and other (8.8%, n = 54). Nearly half of the participants reported never having donated before the survey (n = 531). Interestingly, within the group of never donors, 89.1% (n = 473) expressed a willingness to donate in the future. Among them, 39.5% (n = 210) expressed a commitment to do so regularly (at least once per year), 26.7% (n = 142) would consider it in emergencies for family and friends, and 22.8% (n = 121) would do so in emergencies regardless of the recipient.

A regression analysis was performed to identify the factors associated with blood donation in Guatemala (Table 2). The findings revealed that being female was associated with a 58% lower likelihood of blood donation (aOR:0.42 [0.32,0.55]; p < 0.001). Older age also emerged as having a significant association with donation, as the highest odds of previous donation were observed in those aged 60 or older (aOR: 3.59 [2.14, 6.04]; p < 0.001)]. Additionally, individuals with higher education levels (Master’s or Doctorate) demonstrated a higher rate of previous blood donation (aOR: 1.52 [1.03, 2.24]; p = 0.035). Respondents who knew where to donate and those expressing willingness to donate were more likely to have donated blood (aOR: 2.67 (1.96 - 3.67); p < 0.001 and aOR: 3.74 (2.25-6.21); p < 0.001, respectively).

**Table 2.** Univariate and Multivariate Regression Analyses to identify the factors associated with Blood Donation.

|  | Unadjusted OR<br>(95% CI) | p-value | Adjusted OR<br>(95% CI) | p-value |
| --- | --- | --- | --- | --- |
| **Gender**<br>Male<br>Female | Reference<br>0.53 (0.41 - 0.67) | **<0.001** | 0.42 (0.32 - 0.55) | **<0.001** |
| **Age**<br>19-29<br>30-39<br>40-49<br>50-59<br>60+ | Reference<br>1.05 (0.78-1.40)<br>1.38 (0.95 - 2.01)<br>1.53 (1.03 - 2.27)<br>2.59 (1.62 - 4.15) | 0.757<br>0.087<br>**0.034**<br>**<0.001** | 1.07 (0.77-1.49)<br>1.57 (1.05-2.35)<br>1.76 (1.14-2.71)<br>3.59 (2.14-6.04) | **0.668**<br>**0.029**<br>**0.010**<br>**<0.001** |
| **Educational level**<br>Elementary School /High School<br>Bachelor<br>Master / Doctorate | Reference<br>1.03 (0.74 - 1.43)<br>1.62 (1.14 - 2.29) | 0.867<br>**0.007** | 0.98 (0.69-1.41)<br>1.52 (1.03-2.24) | 0.934<br>**0.035** |
| **Place of residence**<br>Guatemala City<br>Other | Reference<br>1.04 (0.64 - 1.70) | 0.871 |  |  |
| **Do you know where you can donate blood in Guatemala?**<br>No<br>Yes | Reference<br>2.64 (1.97 - 3.54) | **<0.001** | 2.67 (1.96 - 3.67) | **<0.001** |
| **If you were to donate blood, do you care who receives your blood?**<br>No<br>Yes | Reference<br>0.67 (0.47 - 0.97) | **0.033** | 0.94 (0.62 - 1.42) | 0.756 |
| **Are you willing to donate blood?**<br>I am not interested in donating blood<br>Yes, but only for emergencies – it does not matter who receives the blood<br>Yes, but only for emergencies of family, friends, and people I know<br>Yes, I would like to be a frequent donor (1 time per year) | Reference<br>1.84 (1.13-3.01)<br>1.62 (1.00-2.62)<br>2.37 (1.50-3.74) | **<0.001**<br>**0.048**<br>**<0.001** | 2.31 (1.37-3.91)<br>2.13 (1.27-3.61)<br>3.74 (2.25-6.21) | **0.002**<br>**0.004**<br>**<0.001** |

### Barriers and facilitators to blood donation

A total of 1120 respondents provided insights on factors that would facilitate or motivate them to donate blood. Among both never donors and previous donors, improved access to donation centers, specifically an increased number of facilities, which are easily accessible and have extended hours of operation, emerged as the most cited facilitator (Figure 1). The second most frequently mentioned facilitator was improved access to information about blood donation, including details about the procedure, duration, pre- and post-procedure requirements, recovery time, and side effects. Despite these challenges, respondents indicated an altruistic desire to help others as the strongest motivator for donation. Additional challenges identified from survey responses included the need for a more efficient and expedited donation process, increased information on how donated blood is utilized, a lack of incentives for donation, the need for attentive and respectful staff at donation centers, as well as logistical assistance coordinating appointments and reminders for future donations.

Among those who had never donated, the most cited barriers to blood donation included medical contraindications (especially age restrictions and history of transfusion-transmissible infections), unwillingness to donate blood to strangers, and limited operating hours of donation centers. Additional reported barriers can be found in Figure 2. A smaller subgroup also expressed hesitancy regarding the proper utilization of donated blood, expressing apprehension about potential illegal sales or unequal distribution among the population.

**Figure 1.**
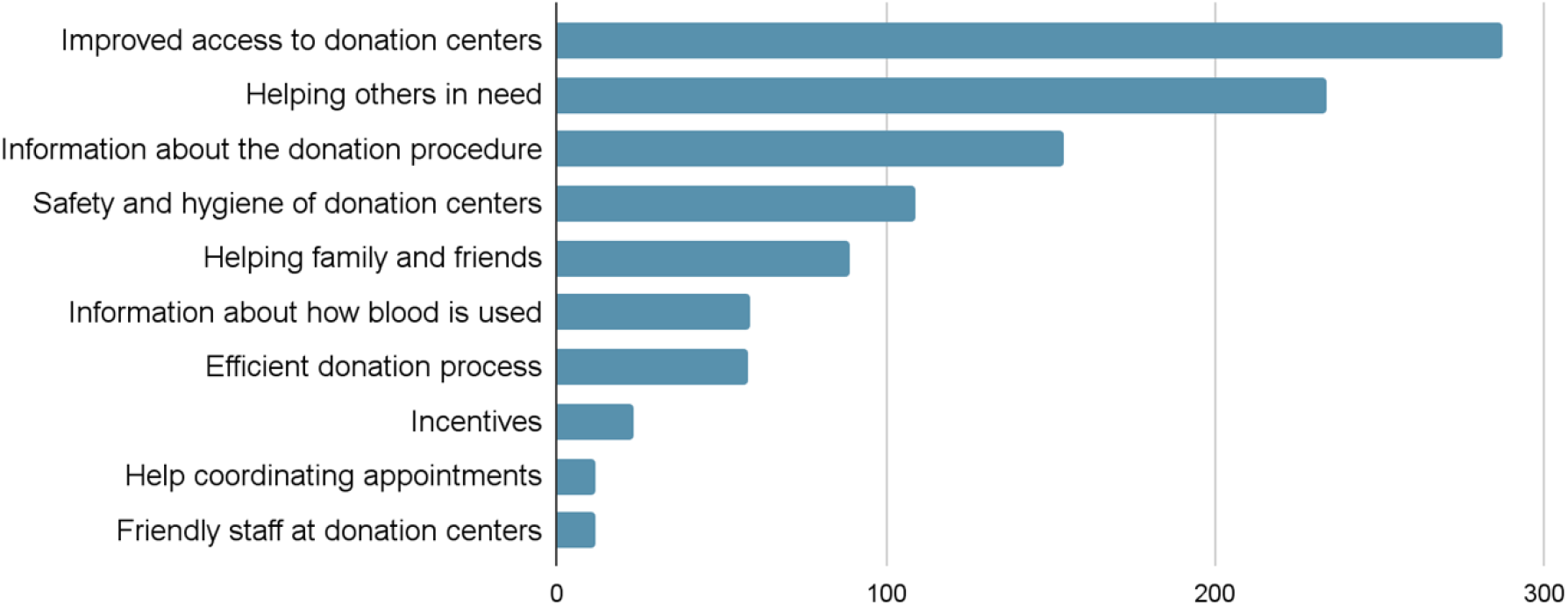
Most common facilitators and motivators for blood donation (n = 1120)

**Figure 2.**
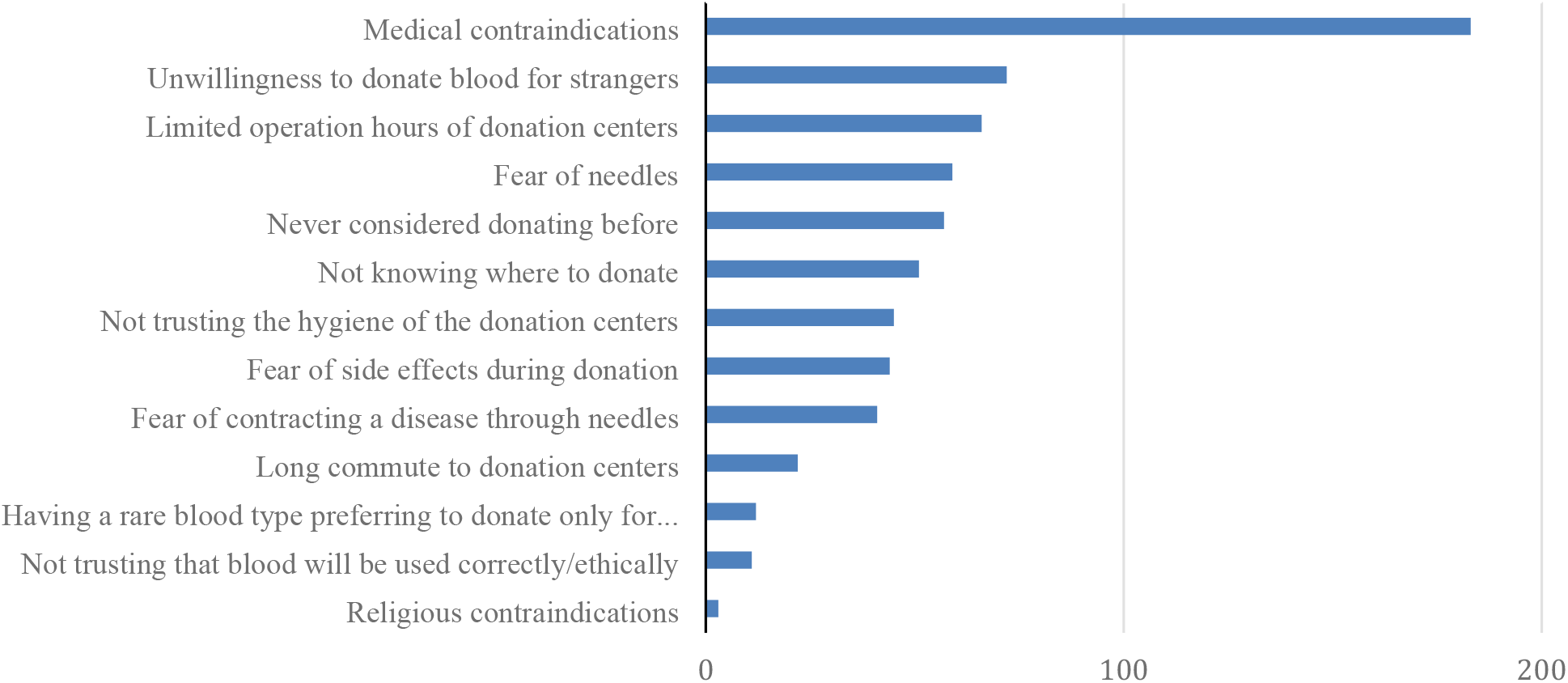
Most common barriers to donation among never donors (n = 531)

## DISCUSSION

The availability of a safe and reliable supply of blood products is a cornerstone and a benchmark of the efficacy of a nation’s healthcare system. These products serve a vital role in a wide range of medical scenarios, from critical interventions in severe trauma or childbirth to supporting patients with hematologic disorders and chronic illnesses. Globally, the availability of blood products relies heavily on voluntary contributions from altruistic donors. This challenge is paramount in a country like Guatemala, grappling with one of the region’s lowest voluntary blood donation rates. The ramifications of this issue extend across various dimensions of the healthcare system, which often lacks the infrastructure and coordination to maximize the limited availability of products. In addition, it is essential to explore the underlying challenges of the population to integrate viable strategies for improvement.

Notably, the COVID-19 pandemic has exacerbated the existing shortage of voluntary blood donations in Latin America and the Caribbean. In Guatemala, the decrease from 4.1% to 3.7% of voluntary donors during 2020 underscores the vulnerability of the blood supply chain in times of crisis and the increased urgency to fortify initiatives and strategies to bolster voluntary blood donation, not only as a response to the current pandemic but also in preparation for potential future health emergencies.^9,10^

Despite the willingness to donate blood expressed by over 90% of respondents in this study, various barriers contributing to the low voluntary donation rate were identified. The primary barrier, limited access to donation centers, presents a multifaceted challenge rooted in geographical disparities, transportation constraints, and restricted operating hours. With only 37 transfusional centers serving a population exceeding 17 million inhabitants, addressing this issue is critical for the Guatemalan population. Additionally, the lack of public funding and resources and the absence of a coordinated national blood blank system hinder the efficient collection and distribution of blood products between centers.^11^ Rectifying the inefficient management and conservation practices of blood products is also integral to optimizing blood transfusion practices. Improvements in inventory control, expiration monitoring, and timely utilization protocols are critical to improving quality indicators such as the discard rate of blood components, which can be as high as 30% in Guatemala.^6^ Overcoming these challenges necessitates a strategic approach involving investment in infrastructure and human resources but has critical potential to help improve the culture of voluntary blood donation in the country. For example, securing the timely utilization of donated products will maximize the impact of voluntary blood donations and address population concerns about the correct utilization of products, particularly in regions affected by violence and corruption. The increased availability of donation opportunities can also assist in counteracting the current inefficient donation process characterized by extended wait times, inattentive staff, and difficulties in scheduling appointments.

The challenges identified in Guatemala are similar to those encountered in other low- and middle-income countries.^12–16^ Particularly in Latin America, countries like Argentina, Costa Rica, Chile, and Nicaragua have implemented successful strategies to improve the availability of blood products, shifting from a replacement-based model to an altruistic donation model, primarily through the reorganization of the structure of the blood collection system. The implementation of the Regional Plan of Action for Transfusion Safety developed by the PAHO, which strategies include the planning and management of the national blood network system, the promotion of voluntary blood donation, the quality assurance, and the appropriate use of blood and blood components, helped increase the voluntary blood donation rate from 38% to 100% in Nicaragua.^17^ Chile increased altruistic donation from 8% to 22% by reorganizing autonomously functioning blood collection centers into transfusion centers, modernizing the data collection systems, and implementing mobile blood collection campaigns.^18,19^ Mobile blood collection units have proven successful in increasing voluntary donation rates in other LMICs and hold promise as a solution to this multifaceted barrier.^20^ These initiatives will require the commitment and leadership of health authorities to integrate these improvements into the national health system.^21^ The government’s commitment to recognizing the importance of blood transfusion and supporting the health care system with the appropriate infrastructural, human, and financial resources is indispensable for achieving these goals.^22^

The request by survey respondents to increase education around voluntary blood donation presents an excellent opportunity to establish partnerships between the government and civil societies. Targeted educational campaigns are critical to promoting a positive culture of voluntary blood donation in the country. Community engagement efforts become crucial in establishing trust and confidence among potential donors by dispelling misconceptions and fears surrounding blood donation and providing accurate information about the safety, risks, and impact of blood donation. The design of culturally sensitive and demographically tailored approaches can have a positive impact on the population.^23,24^ Furthermore, creating a safe and welcoming atmosphere where donors feel at ease and confident that their requirements during the donation process are being addressed can encourage a willingness to return for future donations.^25^

Lastly, community engagement and partnerships with governmental and civil organizations, such as NGOs, religious institutions, and societies, are necessary to implement tailored policies that support voluntary donation. Reviewing and updating the national guidelines for donor eligibility and blood safety standards to reflect evidenced-based strategies that support voluntary blood donation is critical to improving the voluntary blood donation rate.^26^ The WHO/PAHO currently recommends a limit age for blood donation that varies between 65 and 81 years.^27^ However, in Guatemala, only those below 55 years of age are allowed to donate blood. The reduction in the interval of time allowed between donations has also proven effective in increasing the blood product supply.^28^ In the post-pandemic era, modifying eligibility criteria for donation emerges as an essential strategy to broaden inclusivity for blood donors. Other nations have implemented this measure through partnerships with civil societies by removing questions related to potential high-risk behaviors during donor screening to remove the stigma associated with these and potentially tap into a broader pool of willing donors.^29^There are important limitations to our study. This electronic survey was distributed through purposive sampling from an NGO (Dona Guate) network, which may have attracted individuals with a higher intention or willingness to donate blood. Additionally, the distribution method of the survey limits our ability to calculate a response rate and impedes the generalizability of the results. Nonetheless, our results are still relevant to the region and can serve as a foundational framework for future research to comprehensively increase voluntary blood donation rates.

## Conclusions

In conclusion, the challenges surrounding voluntary blood donation in Guatemala unveil the imperative for region-specific strategies. These strategies encompass a spectrum of initiatives, from fortifying supply chains to enhancing the accessibility and availability of blood transfusions. We can significantly improve voluntary blood donation rates through concerted efforts, including investment in infrastructure, targeted educational campaigns, and community engagement. Promoting voluntary blood donation addresses the immediate need for blood products and improves healthcare outcomes, thereby advancing public health in the region.

## Data Availability

The datasets generated and/or analyzed during the current study are available from the corresponding author on reasonable request.

## Acknowledgments

The authors would like to acknowledge Dona Guate (https://www.donaguate.com) and their commitment to promoting a positive culture of blood donation in Guatemala.

## Competing Interests

The authors have nothing to disclose.

## Funding Statement

No funding was received for this manuscript.

## Appendix 1

Translated survey instrument in English

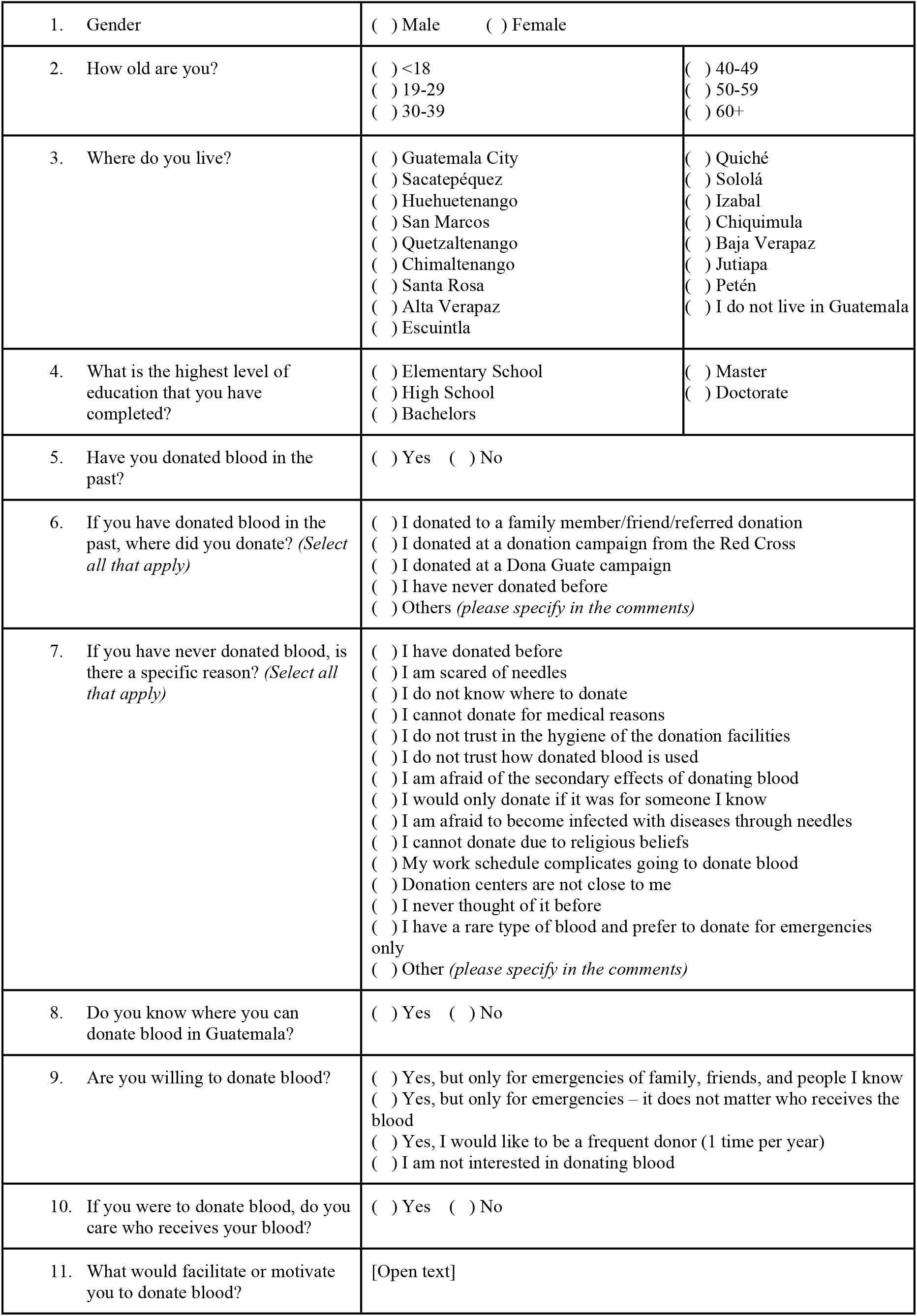

## Notes

### Competing Interest Statement

The authors have declared no competing interest.

### Author Declarations

This research was approved by the Harvard Longwood Campus Institutional Review Board under protocol IRB 23-0135.

